# Association of in-hospital use of ACE-I/ARB and COVID-19 outcomes in African American population

**DOI:** 10.1101/2021.04.14.21255443

**Authors:** Shilong Li, Rangaprasad Sarangarajan, Tomi Jun, Yu-Han Kao, Zichen Wang, Emilio Schadt, Michael A. Kiebish, Elder Granger, Niven R. Narain, Rong Chen, Eric E. Schadt, Li Li

## Abstract

**Importance:** The *ACE* D allele is more prevalent among African Americans (AA) compared to other races/ethnicities and has previously been associated with severe COVID-19 pathogenesis through excessive ACE1 activity. ACE-I/ARBs may counteract this mechanism, but their association with COVID-19 outcomes has not been specifically tested in the AA population.

**Objectives:** To determine whether the use of ACE-I/ARBs is associated with COVID-19 in-hospital mortality in AA compared with non-AA population.

**Design, Setting, and Participants:** In this observational, retrospective study, patient-level data were extracted from the Mount Sinai Health System’s (MSHS) electronic medical record (EMR) database, and 6,218 patients with a laboratory-confirmed COVID-19 diagnosis from February 24 to May 31, 2020 were identified as ACE-I/ARB users.

**Exposures:** Patients with an active prescription from January 1, 2019 up to the date of admission for ACE-I/ARB (outpatient use) and patients administered ACE-I/ARB during hospitalization (in-hospital use) were identified.

**Main Outcomes and Measures:** The primary outcome was in-hospital mortality, assessed in the entire, AA, and non-AA population.

**Results:** Of the 6,218 COVID-19 patients, 1,138 (18.3%) were ACE-I/ARB users. In a multivariate logistic regression model, ACE-I/ARB use was independently associated with reduced risk of in-hospital mortality in the entire population (OR, 0.655; 95% CI, 0.505-0.850; P=0.001), AA population (OR, 0.44; 95% CI, 0.249-0.779; P=0.005), and non-AA population (OR, 0.748, 95% CI, 0.553-1.012, P=0.06). In the AA population, in-hospital use of ACE-I/ARBs was associated with improved mortality (OR, 0.378; 95% CI, 0.188-0.766; P=0.006) while outpatient use was not (OR, 0.889; 95% CI, 0.375-2.158; P=0.812). When analyzing each medication class separately, ARB in-hospital use was significantly associated with reduced in-hospital mortality in the AA population (OR, 0.196; 95% CI, 0.074-0.516; P=0.001), while ACE-I use was not associated with impact on mortality in any population.

**Conclusion and Relevance:** In-hospital use of ARBs was associated with a significant reduction in in-hospital mortality among COVID-19-positive AA patients. These results support further investigation of ARBs to improve outcomes in AA patients at high risk for COVID-19-related mortality.

## Introduction

The current COVID-19 pandemic is expected to reduce USA life expectancy in 2020 by 1.13 years, although life expectancy reductions in African American (AA) and Hispanic populations are estimated to be 3 to 4 times higher than among Caucasians.^1^ While the U.S. African American and Hispanic individuals make up approximately 13.4% and 18.5% of the US population, respectively, they constitute roughly 21% and 22%, respectively, of COVID-19 deaths reported to the National Center for Health Sciences (NCHS).^2^ Socioeconomic factors contribute to these disparities, with 19.7% AA and 16.2% Hispanic individuals, disproportionately represented among essential occupations.^3^ However, it remains unclear whether population differences in the prevalence of genetic risk variants may also contribute to disparate outcomes.

Several studies have attempted to delineate the genetic basis of susceptibility to COVID-19 clinical syndromes.^4–7^ Epidemiological analyses have suggested that polymorphisms in the *ACE* gene, which encodes the angiotensin converting enzyme 1 (ACE1), are related to the incidence and mortality from COVID-19 at the national level.^8–11^ The insertion (I) or deletion (D) of a 287 base pair sequence in intron 16 of the *ACE* gene results in three possible genotypes: DD or II homozygotes, or ID heterozygotes. The D allele has been associated with increased *ACE1* expression in a dose-dependent manner, such that compared to II homozyogtes, ID heterozygotes and DD homozygotes have 31% and 65% more ACE1 protein expression in blood serum, respectively.^12^ Data from predominantly European nations suggests that there is a population-level correlation between the prevalence of the D allele and incidence of COVID-19.^10^

The SARS-CoV-2 virus enters cells by binding to angiotensin-converting enzyme 2 (ACE2). ACE1 and ACE2 have counter-regulatory roles in the renin-angiotensin (RAS) system: ACE1 converts angiotensin I to angiotensin-II, the primary effector of the RAS system, while ACE2 inactivates angiotensin-II. Since SARS-CoV-2 viral binding and entry via ACE2 leads to decreased ACE2 availability, it has been hypothesized that an imbalance in ACE1 and ACE2 activity (leading to excess angiotensin-II) may contribute to the pathogenesis of severe COVID-19.^13^ This imbalance may be further exacerbated by increased *ACE1* expression in the presence of the *ACE* D allele. This mechanistic hypothesis predicts that either ACE1 inhibition or angiotensin-II receptor blockade would have a beneficial effect on COVID-19 outcomes, especially in populations with high prevalence of the *ACE* D allele.^13^

Early in the COVID-19 pandemic, there were concerns that ACE inhibitors (ACE-I) and angiotensin receptor blockers (ARB), commonly used to manage hypertension, might increase *ACE2* expression and thus increase susceptibility to COVID-19. Since then, several groups have assessed the relationship between ACE-I/ARB use and COVID-19 outcomes, with no clear consensus on risk versus benefit.^14–16^ However, these studies were conducted in predominantly Caucasian populations, were focused on estimating risk of COVID-19, and did not account for ethnicity and the *ACE* D allele prevalence. Differences in the frequency of continuing or discontinuing ACE-1/ARB across hospitalized patients may also have contributed to discrepancies between studies.^17^ These results supported the joint statement issued by expert cardiology societies that, in the absence of experimental or clinical information demonstrating benefit or adverse outcomes, individuals using ACE-I/ARB medications should continue their medications during COVID-19 infection.^18^

We recently reported that there was an increased prevalence of the *ACE* D allele among African Americans compared to other races and ethnicities.^13^ We therefore hypothesized that there may be a more pronounced benefit from ACE-I/ARB therapy in the AA population compared to prior published cohorts. This study assessed whether ACE-I/ARB use among hospitalized AA patients with COVID-19 was associated with clinical benefit and improved outcome.

## Methods

### Data source and COVID-19 cohort

This observational, retrospective cohort study utilized deidentified electronic medical record data (EMR data; EPIC systems, Verona, WI) from the Mount Sinai Health System (MSHS). We identified 6,218 patients who met the admission criteria due to COVID-19 admitted to hospital (including emergency room visits and inpatient admissions) related to COVID-19 at a Mount Sinai facility from February 24 through May 31, 2020. Encounters were considered COVID-19-related if a COVID-19 test was ordered or a COVID-19 diagnosis was assigned. COVID-19 was diagnosed by real-time reverse-transcriptase PCR-based clinical tests from nasopharyngeal swab specimens.

We extracted patient demographics from the EMR including age, sex, and self-identified race/ethnicity. Race and ethnicity were summarized into the mutually exclusive categories of African American (AA), Asian, Hispanic, White, Unknown, and Other. We retrieved disease comorbidities from the database including asthma, chronic obstructive pulmonary disease (COPD), obstructive sleep apnea, obesity, diabetes mellitus, chronic kidney disease, HIV infection, cancer, coronary artery disease, atrial fibrillation, heart failure, chronic viral hepatitis, and alcoholic nonalcoholic liver disease. Additionally, we extracted laboratory test orders and results from the encounters including white blood cell (WBC), creatinine, anion gap, potassium and alanine transaminase (ALT). We also retrieved the initial vital sign measurements for each encounter, including BMI, temperature, O_2_ saturation, heart rate, respiratory rate, and blood pressure (BP).

### ACE-I/ARB, other antihypertensive medications and outcomes

We defined ACE-I/ARB exposure as an active prescription from January 1, 2019, up to and including hospitalization for COVID-19 (“anytime use”), with 18.3% in our SARS-CoV-2 positive inpatient population; while 9.4% patients were prescribed ACE-I/ARB in our general population in our EMR system from January 1, 2019 to May 31, 2020. We identified 1,139 anytime use patients and divided them into two subsets: 1) documented ACE-I/ARB administration during their hospitalization (“in-hospital use”, N=605), and 2) ACE-I/ARB use prior to admission but not during hospitalization (“outpatient use”, N=534). We also traced other antihypertensive medications (calcium-channel blockers, beta blockers, and diuretics) dispensed from January 1, 2019 till May 31, 2020, that were included in the medication history or usage. We have listed all the antihypertensive medications in Table S1 in the supplement.

The primary outcome was COVID-19 caused in-hospital mortality in the entire population and in the AA sub-population. There were three possible outcomes for each hospital encounter: in-hospital mortality, discharge, or continued hospitalization (right censored). The secondary outcomes are intensive care unit (ICU) admission and acute kidney injury (AKI), which were evaluated in the AA population as sub-analysis.

We assessed the associations between ACE-I/ARB anytime use, in-hospital use, or outpatient use and COVID-19 in-hospital mortality for the entire population, AA population, or non-AA population compared with patients who were not on ACE-I/ARB. We defined the outpatient use if patients were prescribed ACE-I/ARB during their outpatient visits but without continued in-hospital use, and the in-hospital use if patients continued or were administered ACE-I/ARB. These analyses were also repeated for ACE-I use and ARB use; exposure to either class of medication was determined without regard to exposure to the other class.

### Statistical Analysis

We described continuous variables as their medians and interquartile ranges (IQR), or means and standard deviations (SD). We described categorical variables as a number and percentage (%). We measured statistical differences with Kruskal-Wallis or two sample t-test for continuous variables and Chi-square test for categorical variables, respectively.

We evaluated the ACE-I/ARB associations with in-hospital mortality using multivariate logistic regression adjusting for the following confounders: age, hospital stay duration, race, smoking status, BMI, temperature, O_2_ saturation, heart rate, respiratory rate, hypertension, asthma, chronic obstructive pulmonary disease (COPD), obstructive sleep apnea, obesity, diabetes, chronic kidney disease, HIV, cancer, coronary artery disease, atrial fibrillation, heart failure, chronic viral hepatitis, liver disease, ICU stay, white blood cell (WBC), creatinine, anion gap, potassium and alanine transaminase (ALT). We reported the adjusted odds ratios and corresponding 95% confidence intervals. Statistical significance was defined as a two-sided P<0.05, unless otherwise stated. We also applied the Wald-based power and sample size formulas for logistic regression to calculate the power given the observed sample size.^19^ We used two-tailed test to estimate the probability of event under the null hypothesis which was the in-hospital mortality rate and we used the binomial distribution for the ACE-I/ARB with the prescription rate as the probability. We further analyzed the interaction between ACE-I/ARB with AA race by adding the interaction term in the logistic regression with adjustment by same potential confounders.

We employed inverse propensity-weighted logistic regression as a doubly robust method to balance the potential confounders and further estimate the effect of ACE-I/ARB use on COVID-19 patient outcomes. Specifically, we used the generalized boosted regression to compute propensity scores and balance the covariates based on the average treatment effect on the treated (ATT), and then calculated the adjusted odds ratios and 95% confidence interval using multivariate logistic regression with the balanced covariates. In all the analyses, cases with missing covariates were omitted. We performed all analysis using R (version 4.0.1) and Python (version 3.7). This study is approved by IRB-17-01245.

## Results

### Participants

Our cohort included 6,218 patients diagnosed with COVID-19 and admitted to one of five hospitals using EPIC system in the Mount Sinai Health System (MSHS) from February 24 through May 31, 2020. Among these patients, 1,139 (18.3%) were ACE-I/ARB users and 5,079 (81.7%) were ACE-I/ARB non-users since 2019. The data extraction process and study workflow are shown in Figure 1. ACE-I/ARB users were older than non-users (median age, 68 [IQR, 60-78] vs 60 [IQR, 43-72]) (Table 1). Compared with non-users, ACE-I/ARB users were more likely to have cardiovascular comorbidities and risk factors such as hypertension (60.6% vs 18.3%; P<0.001), diabetes mellitus (40.1% vs 11.3%; P<0.001), coronary artery disease (24.0% vs 5.7%; P<0.001), and heart failure (16.3% vs 3.0%; P<0.001). ACE-I/ARBs users were also more likely to use other antihypertensive medications such as calcium channel blockers (53.8% vs 18.4%; P<0.001), beta blockers (47.7% vs 18.2%; P<0.001), and diuretics (56.4% vs 18.9%; P<0.001). As of May 31, 2020, of the 6,218 patients in our COVID-19 cohort, 1,326 (21.3%) died, 4,384 (70.5%) were discharged, and 508 (8.2%) were still hospitalized as of 31 May 2020.

**Table 1.**
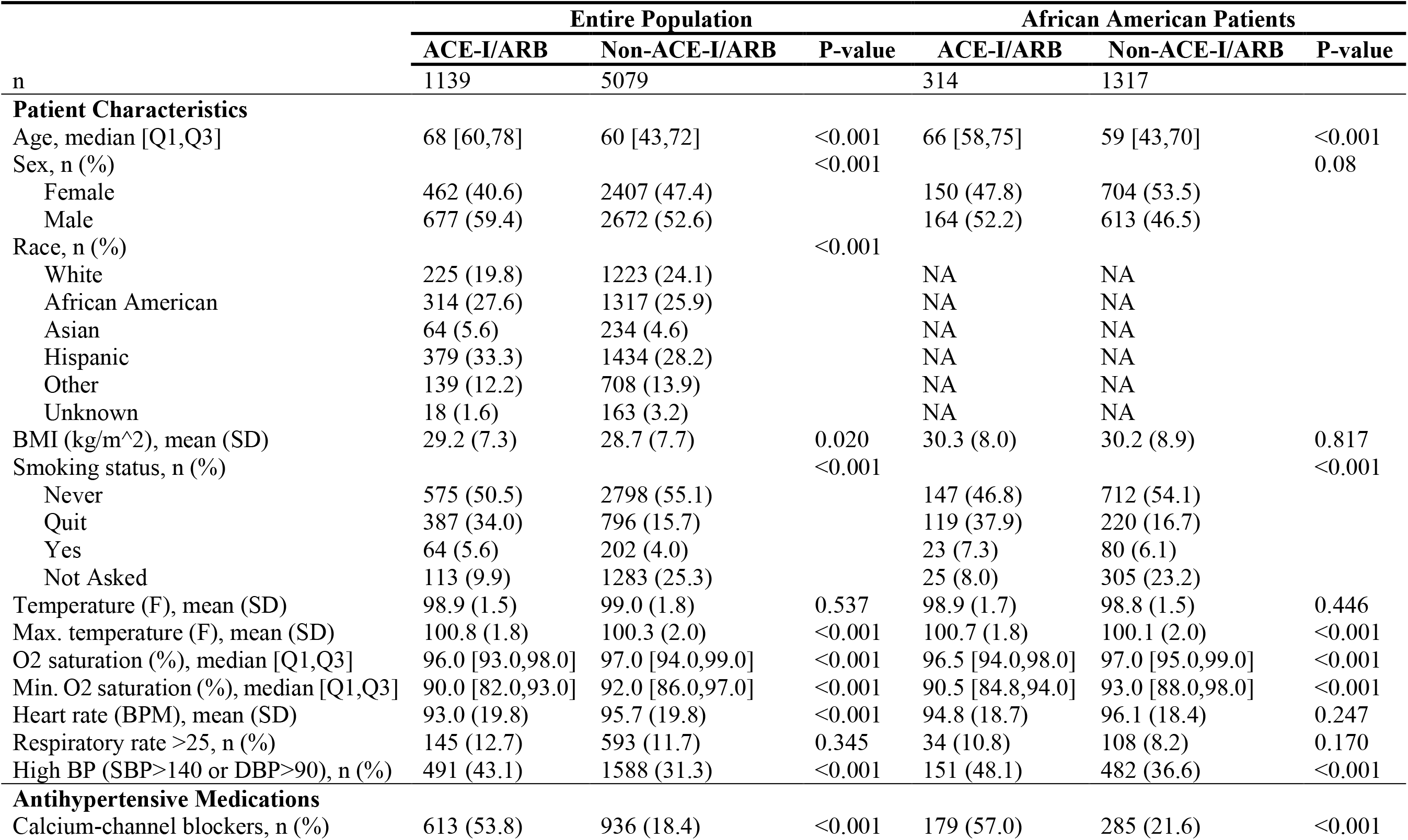
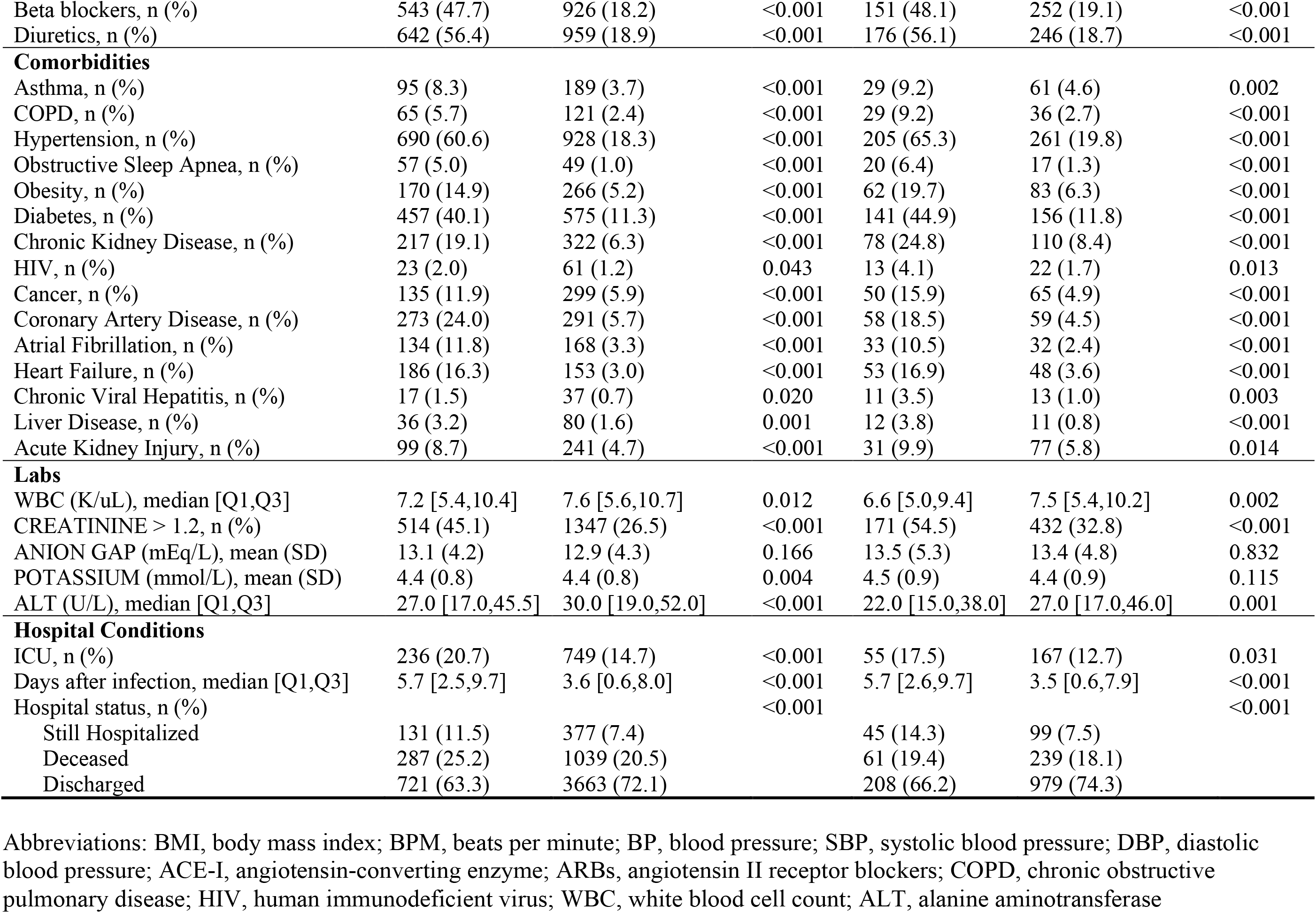
Characteristics of COVID-19 patients in ACE-I/ARB and Non-ACE-I/ARB groups for entire population and African American patients.

|  | Entire Population |  |  | African American Patients |  |  |
| --- | --- | --- | --- | --- | --- | --- |
|  | ACE-I/ARB | Non-ACE-I/ARB | P-value | ACE-I/ARB | Non-ACE-I/ARB | P-value |
| n | 1139 | 5079 |  | 314 | 1317 |  |
| **Patient Characteristics** |  |  |  |  |  |  |
| Age, median [Q1,Q3] | 68 [60,78] | 60 [43,72] | <0.001 | 66 [58,75] | 59 [43,70] | <0.001 |
| Sex, n (%) |  |  | <0.001 |  |  | 0.08 |
| Female | 462 (40.6) | 2407 (47.4) |  | 150 (47.8) | 704 (53.5) |  |
| Male | 677 (59.4) | 2672 (52.6) |  | 164 (52.2) | 613 (46.5) |  |
| Race, n (%) |  |  | <0.001 |  |  |  |
| White | 225 (19.8) | 1223 (24.1) |  | NA | NA |  |
| African American | 314 (27.6) | 1317 (25.9) |  | NA | NA |  |
| Asian | 64 (5.6) | 234 (4.6) |  | NA | NA |  |
| Hispanic | 379 (33.3) | 1434 (28.2) |  | NA | NA |  |
| Other | 139 (12.2) | 708 (13.9) |  | NA | NA |  |
| Unknown | 18 (1.6) | 163 (3.2) |  | NA | NA |  |
| BMI (kg/m<sup>2</sup>), mean (SD) | 29.2 (7.3) | 28.7 (7.7) | 0.020 | 30.3 (8.0) | 30.2 (8.9) | 0.817 |
| Smoking status, n (%) |  |  | <0.001 |  |  | <0.001 |
| Never | 575 (50.5) | 2798 (55.1) |  | 147 (46.8) | 712 (54.1) |  |
| Quit | 387 (34.0) | 796 (15.7) |  | 119 (37.9) | 220 (16.7) |  |
| Yes | 64 (5.6) | 202 (4.0) |  | 23 (7.3) | 80 (6.1) |  |
| Not Asked | 113 (9.9) | 1283 (25.3) |  | 25 (8.0) | 305 (23.2) |  |
| Temperature (F), mean (SD) | 98.9 (1.5) | 99.0 (1.8) | 0.537 | 98.9 (1.7) | 98.8 (1.5) | 0.446 |
| Max. temperature (F), mean (SD) | 100.8 (1.8) | 100.3 (2.0) | <0.001 | 100.7 (1.8) | 100.1 (2.0) | <0.001 |
| O2 saturation (%), median [Q1,Q3] | 96.0 [93.0,98.0] | 97.0 [94.0,99.0] | <0.001 | 96.5 [94.0,98.0] | 97.0 [95.0,99.0] | <0.001 |
| Min. O2 saturation (%), median [Q1,Q3] | 90.0 [82.0,93.0] | 92.0 [86.0,97.0] | <0.001 | 90.5 [84.8,94.0] | 93.0 [88.0,98.0] | <0.001 |
| Heart rate (BPM), mean (SD) | 93.0 (19.8) | 95.7 (19.8) | <0.001 | 94.8 (18.7) | 96.1 (18.4) | 0.247 |
| Respiratory rate >25, n (%) | 145 (12.7) | 593 (11.7) | 0.345 | 34 (10.8) | 108 (8.2) | 0.170 |
| High BP (SBP>140 or DBP>90), n (%) | 491 (43.1) | 1588 (31.3) | <0.001 | 151 (48.1) | 482 (36.6) | <0.001 |
| **Antihypertensive Medications** |  |  |  |  |  |  |
| Calcium-channel blockers, n (%) | 613 (53.8) | 936 (18.4) | <0.001 | 179 (57.0) | 285 (21.6) | <0.001 |
| Beta blockers, n (%) | 543 (47.7) | 926 (18.2) | <0.001 | 151 (48.1) | 252 (19.1) | <0.001 |
| Diuretics, n (%) | 642 (56.4) | 959 (18.9) | <0.001 | 176 (56.1) | 246 (18.7) | <0.001 |
| **Comorbidities** |  |  |  |  |  |  |
| Asthma, n (%) | 95 (8.3) | 189 (3.7) | <0.001 | 29 (9.2) | 61 (4.6) | 0.002 |
| COPD, n (%) | 65 (5.7) | 121 (2.4) | <0.001 | 29 (9.2) | 36 (2.7) | <0.001 |
| Hypertension, n (%) | 690 (60.6) | 928 (18.3) | <0.001 | 205 (65.3) | 261 (19.8) | <0.001 |
| Obstructive Sleep Apnea, n (%) | 57 (5.0) | 49 (1.0) | <0.001 | 20 (6.4) | 17 (1.3) | <0.001 |
| Obesity, n (%) | 170 (14.9) | 266 (5.2) | <0.001 | 62 (19.7) | 83 (6.3) | <0.001 |
| Diabetes, n (%) | 457 (40.1) | 575 (11.3) | <0.001 | 141 (44.9) | 156 (11.8) | <0.001 |
| Chronic Kidney Disease, n (%) | 217 (19.1) | 322 (6.3) | <0.001 | 78 (24.8) | 110 (8.4) | <0.001 |
| HIV, n (%) | 23 (2.0) | 61 (1.2) | 0.043 | 13 (4.1) | 22 (1.7) | 0.013 |
| Cancer, n (%) | 135 (11.9) | 299 (5.9) | <0.001 | 50 (15.9) | 65 (4.9) | <0.001 |
| Coronary Artery Disease, n (%) | 273 (24.0) | 291 (5.7) | <0.001 | 58 (18.5) | 59 (4.5) | <0.001 |
| Atrial Fibrillation, n (%) | 134 (11.8) | 168 (3.3) | <0.001 | 33 (10.5) | 32 (2.4) | <0.001 |
| Heart Failure, n (%) | 186 (16.3) | 153 (3.0) | <0.001 | 53 (16.9) | 48 (3.6) | <0.001 |
| Chronic Viral Hepatitis, n (%) | 17 (1.5) | 37 (0.7) | 0.020 | 11 (3.5) | 13 (1.0) | 0.003 |
| Liver Disease, n (%) | 36 (3.2) | 80 (1.6) | 0.001 | 12 (3.8) | 11 (0.8) | <0.001 |
| Acute Kidney Injury, n (%) | 99 (8.7) | 241 (4.7) | <0.001 | 31 (9.9) | 77 (5.8) | 0.014 |
| **Labs** |  |  |  |  |  |  |
| WBC (K/uL), median [Q1,Q3] | 7.2 [5.4,10.4] | 7.6 [5.6,10.7] | 0.012 | 6.6 [5.0,9.4] | 7.5 [5.4,10.2] | 0.002 |
| CREATININE > 1.2, n (%) | 514 (45.1) | 1347 (26.5) | <0.001 | 171 (54.5) | 432 (32.8) | <0.001 |
| ANION GAP (mEq/L), mean (SD) | 13.1 (4.2) | 12.9 (4.3) | 0.166 | 13.5 (5.3) | 13.4 (4.8) | 0.832 |
| POTASSIUM (mmol/L), mean (SD) | 4.4 (0.8) | 4.4 (0.8) | 0.004 | 4.5 (0.9) | 4.4 (0.9) | 0.115 |
| ALT (U/L), median [Q1,Q3] | 27.0 [17.0,45.5] | 30.0 [19.0,52.0] | <0.001 | 22.0 [15.0,38.0] | 27.0 [17.0,46.0] | 0.001 |
| **Hospital Conditions** |  |  |  |  |  |  |
| ICU, n (%) | 236 (20.7) | 749 (14.7) | <0.001 | 55 (17.5) | 167 (12.7) | 0.031 |
| Days after infection, median [Q1,Q3] | 5.7 [2.5,9.7] | 3.6 [0.6,8.0] | <0.001 | 5.7 [2.6,9.7] | 3.5 [0.6,7.9] | <0.001 |
| Hospital status, n (%) |  |  | <0.001 |  |  | <0.001 |
| Still Hospitalized | 131 (11.5) | 377 (7.4) |  | 45 (14.3) | 99 (7.5) |  |
| Deceased | 287 (25.2) | 1039 (20.5) |  | 61 (19.4) | 239 (18.1) |  |
| Discharged | 721 (63.3) | 3663 (72.1) |  | 208 (66.2) | 979 (74.3) |  |

Abbreviations: BMI, body mass index; BPM, beats per minute; BP, blood pressure; SBP, systolic blood pressure; DBP, diastolic blood pressure; ACE-I, angiotensin-converting enzyme; ARBs, angiotensin II receptor blockers; COPD, chronic obstructive pulmonary disease; HIV, human immunodeficient virus; WBC, white blood cell count; ALT, alanine aminotransferase

**Figure 1.**
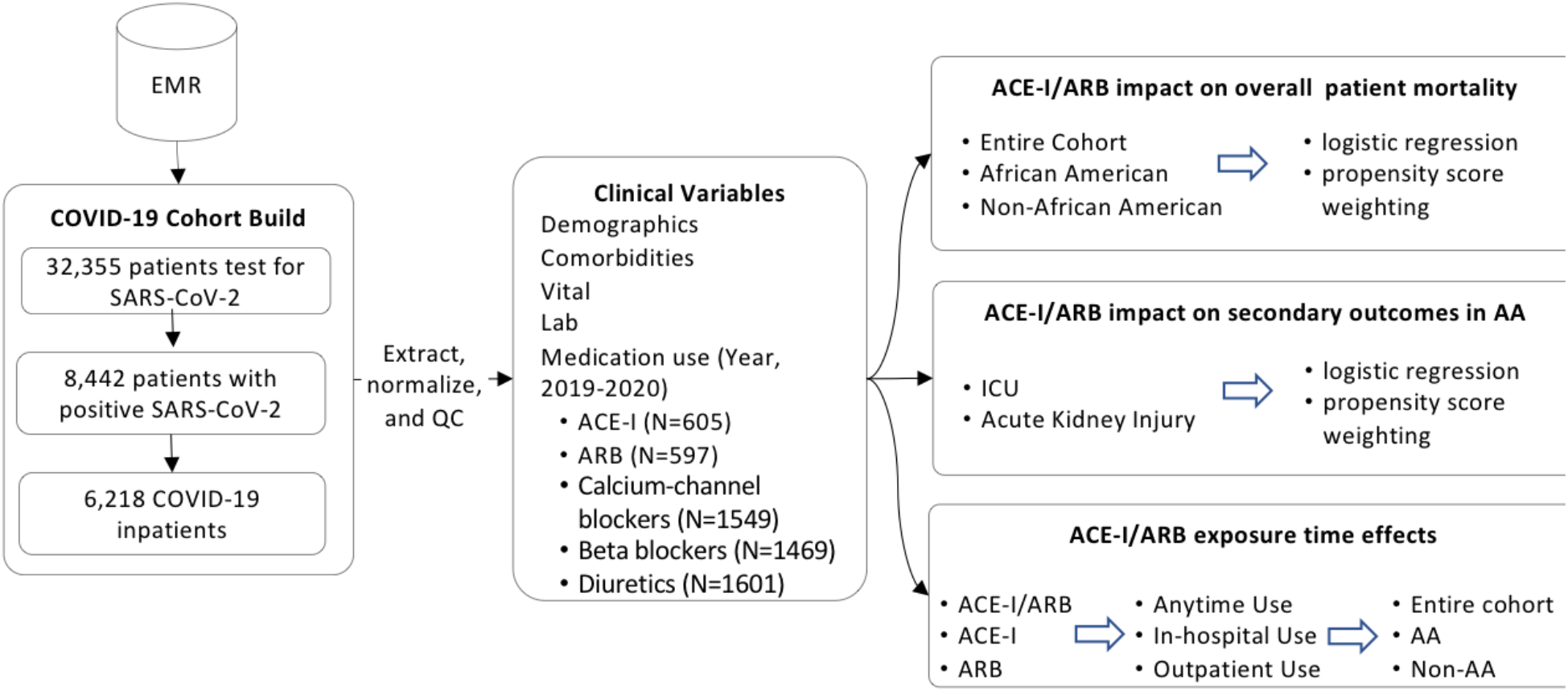
The working flowchart for analyzing ACE-I/ARB effects on COVID-19 in-hospital mortality, ICU admission, and AKI development for African American population. Abbreviations: EMR: electronic medical records; QC, quality control; ACE-I, angiotensin-converting enzyme inhibitor; ARB, angiotensin receptor blocker; ICU, intensive care unit; AA, African American

African American (AA) patients comprised 26.2% of our COVID-19 cohort (1,621 patients). Similar to the entire population, AA ACE-I/ARB users were older than ACE-I/ARB non-users (median age, 66 [IQR 58-75] vs 59 [IQR 43-70]) and more likely to have disease comorbidities (Table 1). In the AA population, 300 patients (18.4%) died, 1,187 patients (72.8%) were discharged, and 114 patients (8.8%) were still hospitalized as of May 31, 2020. A comparison of the characteristics of the AA and non-AA populations is presented in Table S2

### ACE-I/ARB associations with in-hospital mortality overall and by race

Unadjusted mortality was higher in ACE-I/ARB users compared to non-users in both the entire population (25.2% vs. 20.5%, P<0.001) and AA population (19.4% vs. 18.1%, P=0.296) (Table 1), especially for hypertension, obesity, and diabetes, which are known risk factors to COVID mortality.^20^ We therefore adjusted for these confounding factors in this population substantially enriched with chronic conditions and observed that ACE-I/ARB use was independently associated with reduced risk of in-hospital mortality in the entire population (OR, 0.655; 95% CI, 0.505-0.850; P=0.001) by the multivariable logistic regression model (Table 2). Although the interaction term of ACE-I/ARB use and AA population is not significant (OR, 0.736; 95% CI, 0.428-1.266; P=0.268), the odds of in-hospital death in AA population who received ACE-I/ARB is smaller compared to non-AA population who also received ACE-I/ARB (OR, 0.659) (Table S2). When stratified by sub-population, there was a significant reduction in in-hospital mortality in the AA population (OR, 0.44; 95% CI, 0.249-0.779; P=0.005), and did not reach significance in the non-AA population (OR, 0.748; 95% CI, 0.553-1.012; P=0.06) with the power 1-β= 0.8 in the Wald-based power analysis. The prescription ratio of ACE-I/ARB was similar between the AA and non-AA population (Table S3). We confirmed this result using inverse propensity-weighted logistic regression after balancing for confounders: ACE-I/ARB use was significantly associated with decreased risk of in-hospital mortality in the entire population (OR, 0.673; 95% CI, 0.511-0.886; P=0.005), the AA population (OR, 0.381; 95% CI, 0.204-0.712; P=0.003), but not the non-AA population (OR, 0.765; 95% CI, 0.556-1.052; P=0.1)

**Table 2.**
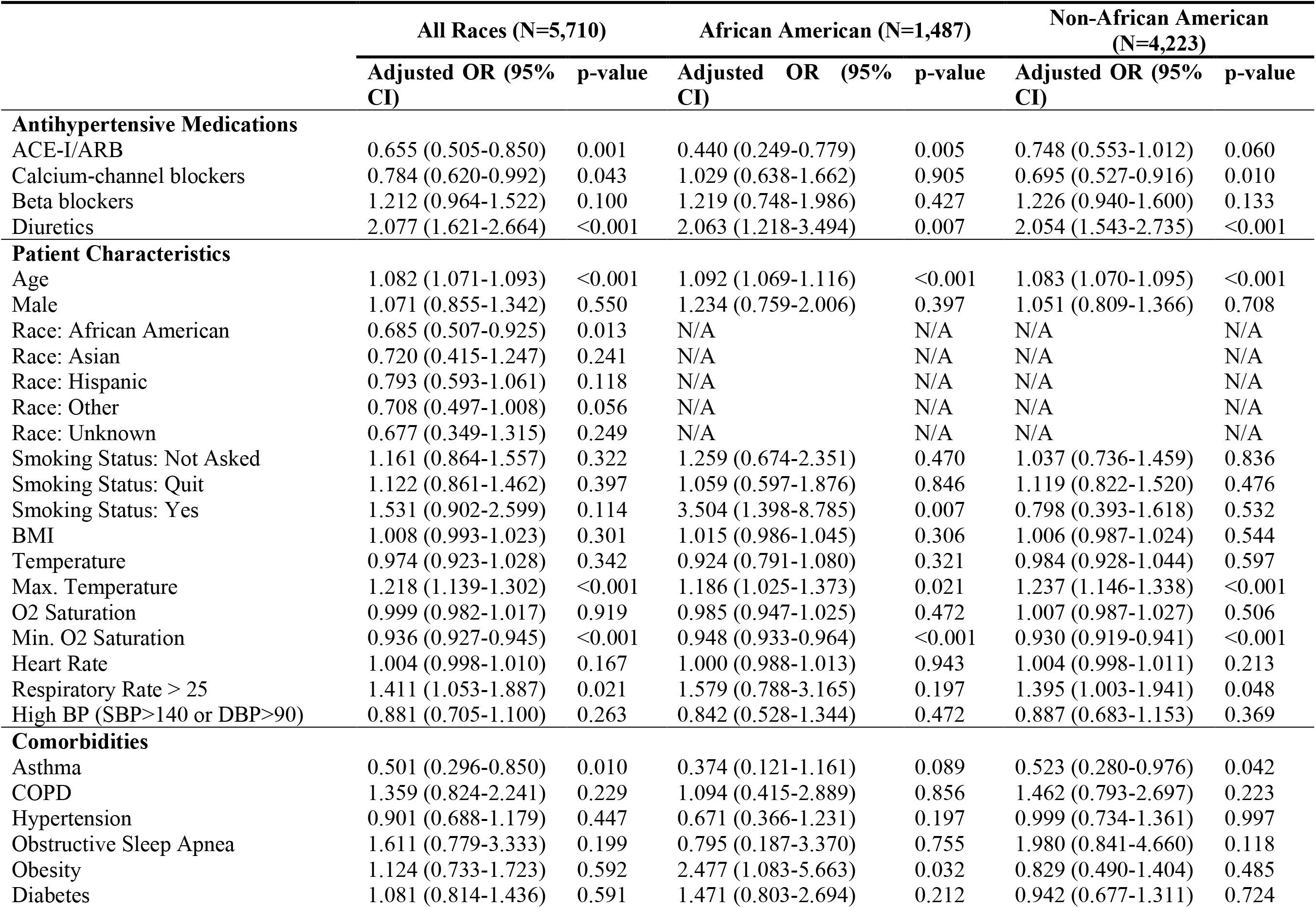
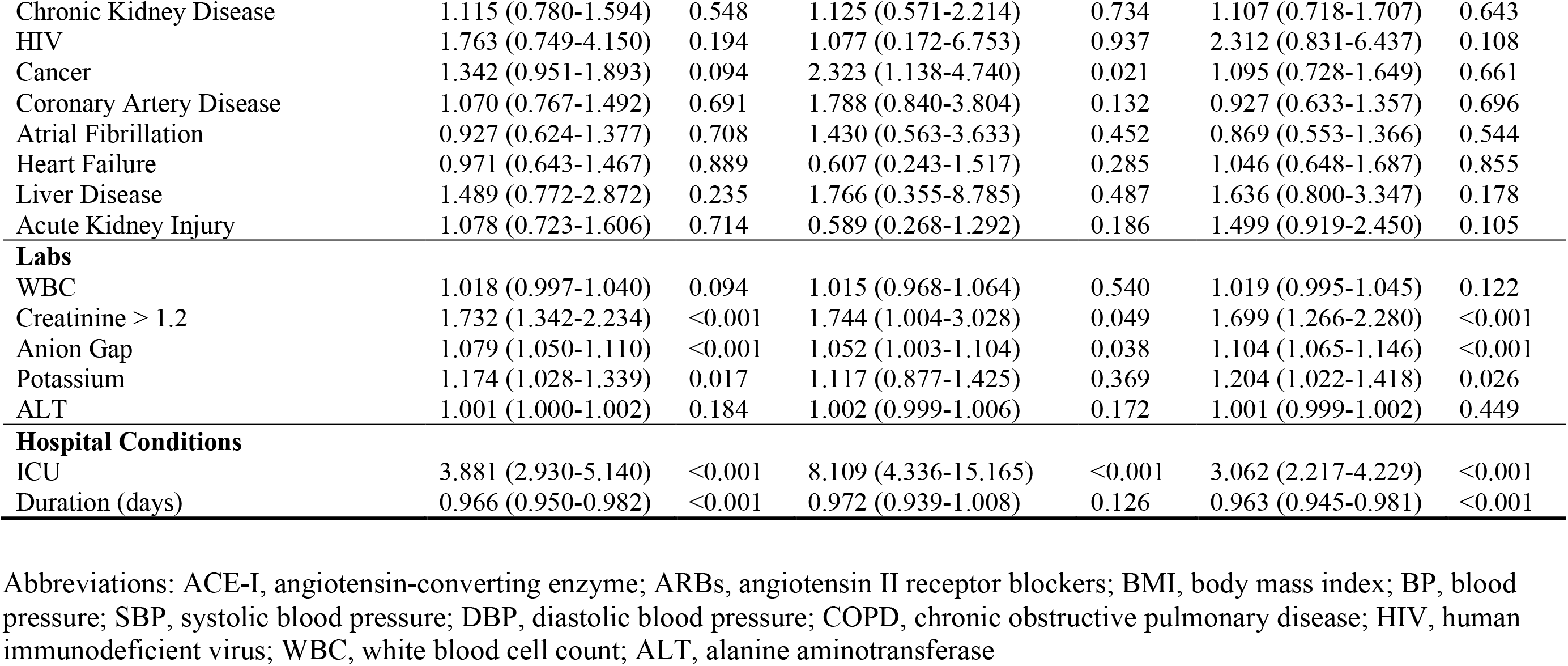
Adjusted odds ratios for COVID-19 in-hospital mortality in different race groups associated with ACE-I/ARB and other covariates.

|  | All Races (N=5,710) |  |  | African American (N=1,487) |  |  | Non-African American (N=4,223) |  |
| --- | --- | --- | --- | --- | --- | --- | --- | --- |
|  | Adjusted OR (95% CI) | p-value |  | Adjusted OR (95% CI) | p-value |  | Adjusted OR (95% CI) | p-value |
| **Antihypertensive Medications** |  |  |  |  |  |  |  |  |
| ACE-I/ARB | 0.655 (0.505-0.850) | 0.001 |  | 0.440 (0.249-0.779) | 0.005 |  | 0.748 (0.553-1.012) | 0.060 |
| Calcium-channel blockers | 0.784 (0.620-0.992) | 0.043 |  | 1.029 (0.638-1.662) | 0.905 |  | 0.695 (0.527-0.916) | 0.010 |
| Beta blockers | 1.212 (0.964-1.522) | 0.100 |  | 1.219 (0.748-1.986) | 0.427 |  | 1.226 (0.940-1.600) | 0.133 |
| Diuretics | 2.077 (1.621-2.664) | <0.001 |  | 2.063 (1.218-3.494) | 0.007 |  | 2.054 (1.543-2.735) | <0.001 |
| **Patient Characteristics** |  |  |  |  |  |  |  |  |
| Age | 1.082 (1.071-1.093) | <0.001 |  | 1.092 (1.069-1.116) | <0.001 |  | 1.083 (1.070-1.095) | <0.001 |
| Male | 1.071 (0.855-1.342) | 0.550 |  | 1.234 (0.759-2.006) | 0.397 |  | 1.051 (0.809-1.366) | 0.708 |
| Race: African American | 0.685 (0.507-0.925) | 0.013 |  | N/A | N/A |  | N/A | N/A |
| Race: Asian | 0.720 (0.415-1.247) | 0.241 |  | N/A | N/A |  | N/A | N/A |
| Race: Hispanic | 0.793 (0.593-1.061) | 0.118 |  | N/A | N/A |  | N/A | N/A |
| Race: Other | 0.708 (0.497-1.008) | 0.056 |  | N/A | N/A |  | N/A | N/A |
| Race: Unknown | 0.677 (0.349-1.315) | 0.249 |  | N/A | N/A |  | N/A | N/A |
| Smoking Status: Not Asked | 1.161 (0.864-1.557) | 0.322 |  | 1.259 (0.674-2.351) | 0.470 |  | 1.037 (0.736-1.459) | 0.836 |
| Smoking Status: Quit | 1.122 (0.861-1.462) | 0.397 |  | 1.059 (0.597-1.876) | 0.846 |  | 1.119 (0.822-1.520) | 0.476 |
| Smoking Status: Yes | 1.531 (0.902-2.599) | 0.114 |  | 3.504 (1.398-8.785) | 0.007 |  | 0.798 (0.393-1.618) | 0.532 |
| BMI | 1.008 (0.993-1.023) | 0.301 |  | 1.015 (0.986-1.045) | 0.306 |  | 1.006 (0.987-1.024) | 0.544 |
| Temperature | 0.974 (0.923-1.028) | 0.342 |  | 0.924 (0.791-1.080) | 0.321 |  | 0.984 (0.928-1.044) | 0.597 |
| Max. Temperature | 1.218 (1.139-1.302) | <0.001 |  | 1.186 (1.025-1.373) | 0.021 |  | 1.237 (1.146-1.338) | <0.001 |
| O2 Saturation | 0.999 (0.982-1.017) | 0.919 |  | 0.985 (0.947-1.025) | 0.472 |  | 1.007 (0.987-1.027) | 0.506 |
| Min. O2 Saturation | 0.936 (0.927-0.945) | <0.001 |  | 0.948 (0.933-0.964) | <0.001 |  | 0.930 (0.919-0.941) | <0.001 |
| Heart Rate | 1.004 (0.998-1.010) | 0.167 |  | 1.000 (0.988-1.013) | 0.943 |  | 1.004 (0.998-1.011) | 0.213 |
| Respiratory Rate > 25 | 1.411 (1.053-1.887) | 0.021 |  | 1.579 (0.788-3.165) | 0.197 |  | 1.395 (1.003-1.941) | 0.048 |
| High BP (SBP>140 or DBP>90) | 0.881 (0.705-1.100) | 0.263 |  | 0.842 (0.528-1.344) | 0.472 |  | 0.887 (0.683-1.153) | 0.369 |
| **Comorbidities** |  |  |  |  |  |  |  |  |
| Asthma | 0.501 (0.296-0.850) | 0.010 |  | 0.374 (0.121-1.161) | 0.089 |  | 0.523 (0.280-0.976) | 0.042 |
| COPD | 1.359 (0.824-2.241) | 0.229 |  | 1.094 (0.415-2.889) | 0.856 |  | 1.462 (0.793-2.697) | 0.223 |
| Hypertension | 0.901 (0.688-1.179) | 0.447 |  | 0.671 (0.366-1.231) | 0.197 |  | 0.999 (0.734-1.361) | 0.997 |
| Obstructive Sleep Apnea | 1.611 (0.779-3.333) | 0.199 |  | 0.795 (0.187-3.370) | 0.755 |  | 1.980 (0.841-4.660) | 0.118 |
| Obesity | 1.124 (0.733-1.723) | 0.592 |  | 2.477 (1.083-5.663) | 0.032 |  | 0.829 (0.490-1.404) | 0.485 |
| Diabetes | 1.081 (0.814-1.436) | 0.591 |  | 1.471 (0.803-2.694) | 0.212 |  | 0.942 (0.677-1.311) | 0.724 |

|  |  |  |  |  |  |  |
| --- | --- | --- | --- | --- | --- | --- |
| Chronic Kidney Disease | 1.115 (0.780-1.594) | 0.548 | 1.125 (0.571-2.214) | 0.734 | 1.107 (0.718-1.707) | 0.643 |
| HIV | 1.763 (0.749-4.150) | 0.194 | 1.077 (0.172-6.753) | 0.937 | 2.312 (0.831-6.437) | 0.108 |
| Cancer | 1.342 (0.951-1.893) | 0.094 | 2.323 (1.138-4.740) | 0.021 | 1.095 (0.728-1.649) | 0.661 |
| Coronary Artery Disease | 1.070 (0.767-1.492) | 0.691 | 1.788 (0.840-3.804) | 0.132 | 0.927 (0.633-1.357) | 0.696 |
| Atrial Fibrillation | 0.927 (0.624-1.377) | 0.708 | 1.430 (0.563-3.633) | 0.452 | 0.869 (0.553-1.366) | 0.544 |
| Heart Failure | 0.971 (0.643-1.467) | 0.889 | 0.607 (0.243-1.517) | 0.285 | 1.046 (0.648-1.687) | 0.855 |
| Liver Disease | 1.489 (0.772-2.872) | 0.235 | 1.766 (0.355-8.785) | 0.487 | 1.636 (0.800-3.347) | 0.178 |
| Acute Kidney Injury | 1.078 (0.723-1.606) | 0.714 | 0.589 (0.268-1.292) | 0.186 | 1.499 (0.919-2.450) | 0.105 |
| **Labs** |  |  |  |  |  |  |
| WBC | 1.018 (0.997-1.040) | 0.094 | 1.015 (0.968-1.064) | 0.540 | 1.019 (0.995-1.045) | 0.122 |
| Creatinine > 1.2 | 1.732 (1.342-2.234) | <0.001 | 1.744 (1.004-3.028) | 0.049 | 1.699 (1.266-2.280) | <0.001 |
| Anion Gap | 1.079 (1.050-1.110) | <0.001 | 1.052 (1.003-1.104) | 0.038 | 1.104 (1.065-1.146) | <0.001 |
| Potassium | 1.174 (1.028-1.339) | 0.017 | 1.117 (0.877-1.425) | 0.369 | 1.204 (1.022-1.418) | 0.026 |
| ALT | 1.001 (1.000-1.002) | 0.184 | 1.002 (0.999-1.006) | 0.172 | 1.001 (0.999-1.002) | 0.449 |
| **Hospital Conditions** |  |  |  |  |  |  |
| ICU | 3.881 (2.930-5.140) | <0.001 | 8.109 (4.336-15.165) | <0.001 | 3.062 (2.217-4.229) | <0.001 |
| Duration (days) | 0.966 (0.950-0.982) | <0.001 | 0.972 (0.939-1.008) | 0.126 | 0.963 (0.945-0.981) | <0.001 |

Abbreviations: ACE-I, angiotensin-converting enzyme; ARBs, angiotensin II receptor blockers; BMI, body mass index; BP, blood pressure; SBP, systolic blood pressure; DBP, diastolic blood pressure; COPD, chronic obstructive pulmonary disease; HIV, human immunodeficient virus; WBC, white blood cell count; ALT, alanine aminotransferase

### ACE-I/ARB use among African Americans did not impact secondary outcomes

In the AA population, since we identified the significant association between ACE-I/ARB use and in-hospital mortality, we further estimated the effects of ACE-I/ARB use on two secondary outcomes: intensive care unit (ICU) admission and acute kidney injury (AKI) development. Unadjusted rates of these outcomes were higher among AA ACE-I/ARB users than AA non-users (Death: 19.4% vs. 18.1%, P=0.296; ICU: 17.5% vs. 12.7%, P=0.03; AKI: 9.9% vs. 5.8%, P=0.01) (Table 1). Neither ICU admission (OR, 0.709; 95% CI, 0.419-1.198; P=0.198) nor AKI (OR, 1.306; 95% CI, 0.721-2.364; P=0.379) were significantly associated with ACE-I/ARB use in the AA population (Table S4). We confirmed these findings using inverse propensity-weighted logistic regression as well, where again, ACE-I/ARB use was not significantly associated with ICU admission (OR, 0.733; 95% CI, 0.425-1.264; P=0.264) or AKI (OR, 1.257; 95% CI, 0.642-2.463; P=0.505).

### In-hospital ARB use reduced risk of COVID-19 caused in-hospital mortality

Given the significant impact ACE-I/ARB exposure had on in-hospital mortality of AA patients admitted for COVID-19, we sought to distinguish between in-hospital use of these classes of drugs and out-patient use, and between the two classes of drugs. ACE-I/ARB in-hospital use was significantly associated with reduced mortality in the entire population (OR, 0.575; 95% CI, 0.419-0.791; P=0.001), the AA population (OR, 0.378; 95% CI, 0.188-0.766; P=0.006), and the non-AA population (OR, 0.652; 95% CI, 0.449-0.946; P=0.024) (Table 3). Outpatient ACE-I/ARB use alone was not significantly associated with mortality in the entire population (OR, 1.016; 95% CI, 0.69-1.496; P=0.936), the AA population (OR, 0.889; 95% CI, 0.375-2.158; P=0.812), nor the non-AA population (OR, 1.101; 95% CI, 0.7-1.73; P=0.677).

**Table 3.** ACE-I/ARB in-hospital use: adjusted odds ratios for COVID-19 in-hospital mortality in different race groups.

|  | All Races (N=5,240) |  | African American (N=1,356) |  | Non-African American (N=3,884) |  |
| --- | --- | --- | --- | --- | --- | --- |
|  | Adjusted OR (95% CI) | p-value | Adjusted OR (95% CI) | p-value | Adjusted OR (95% CI) | p-value |
| **Antihypertensive Medications** |  |  |  |  |  |  |
| ACE-I/ARB | 0.575 (0.419-0.791) | 0.001 | 0.378 (0.188-0.760) | 0.006 | 0.652 (0.449-0.946) | 0.024 |
| Calcium-channel blockers | 0.745 (0.577-0.964) | 0.025 | 0.919 (0.549-1.540) | 0.75 | 0.678 (0.501-0.919) | 0.012 |
| Beta blockers | 1.218 (0.951-1.560) | 0.118 | 1.229 (0.728-2.071) | 0.44 | 1.265 (0.946-1.690) | 0.113 |
| Diuretics | 1.908 (1.456-2.497) | <0.001 | 1.775 (1.007-3.130) | 0.047 | 1.887 (1.379-2.581) | <0.001 |
| **Patient Characteristics** |  |  |  |  |  |  |
| Age | 1.082 (1.071-1.094) | <0.001 | 1.094 (1.069-1.120) | <0.001 | 1.082 (1.069-1.096) | <0.001 |
| Male | 0.944 (0.740-1.203) | 0.639 | 1.178 (0.698-1.990) | 0.539 | 0.908 (0.684-1.206) | 0.505 |
| Race: African American | 0.755 (0.548-1.040) | 0.085 | N/A | N/A | N/A | N/A |
| Race: Asian | 0.835 (0.463-1.508) | 0.551 | N/A | N/A | N/A | N/A |
| Race: Hispanic | 0.839 (0.614-1.148) | 0.273 | N/A | N/A | N/A | N/A |
| Race: Other | 0.753 (0.516-1.097) | 0.14 | N/A | N/A | N/A | N/A |
| Race: Unknown | 0.740 (0.375-1.459) | 0.385 | N/A | N/A | N/A | N/A |
| Smoking Status: Not Asked | 1.203 (0.888-1.631) | 0.233 | 1.420 (0.746-2.705) | 0.286 | 1.067 (0.749-1.520) | 0.718 |
| Smoking Status: Quit | 1.201 (0.898-1.608) | 0.218 | 1.335 (0.715-2.494) | 0.365 | 1.135 (0.808-1.595) | 0.463 |
| Smoking Status: Yes | 1.624 (0.924-2.852) | 0.092 | 3.963 (1.514-10.360) | 0.005 | 0.757 (0.351-1.631) | 0.476 |
| BMI | 1.008 (0.992-1.024) | 0.329 | 1.015 (0.984-1.046) | 0.352 | 1.009 (0.989-1.028) | 0.394 |
| Temperature | 0.983 (0.929-1.040) | 0.548 | 0.946 (0.803-1.117) | 0.518 | 0.988 (0.929-1.051) | 0.709 |
| Max. Temperature | 1.235 (1.150-1.326) | <0.001 | 1.206 (1.034-1.406) | 0.017 | 1.255 (1.155-1.363) | <0.001 |
| O2 Saturation | 0.992 (0.973-1.011) | 0.409 | 0.987 (0.946-1.028) | 0.533 | 0.997 (0.975-1.019) | 0.784 |
| Min. O2 Saturation | 0.937 (0.927-0.946) | <0.001 | 0.950 (0.934-0.966) | <0.001 | 0.931 (0.919-0.943) | <0.001 |
| Heart Rate | 1.003 (0.997-1.010) | 0.284 | 0.998 (0.985-1.012) | 0.818 | 1.004 (0.996-1.011) | 0.325 |
| Respiratory Rate > 25 | 1.339 (0.977-1.833) | 0.069 | 1.336 (0.619-2.883) | 0.461 | 1.339 (0.941-1.904) | 0.105 |
| High BP (SBP>140 or DBP>90) | 0.873 (0.687-1.108) | 0.265 | 0.787 (0.477-1.298) | 0.348 | 0.868 (0.654-1.150) | 0.325 |
| **Comorbidities** |  |  |  |  |  |  |
| Asthma | 0.548 (0.304-0.989) | 0.046 | 0.222 (0.051-0.965) | 0.045 | 0.650 (0.330-1.281) | 0.213 |
| COPD | 1.575 (0.893-2.779) | 0.117 | 1.793 (0.586-5.490) | 0.307 | 1.610 (0.804-3.222) | 0.179 |
| Hypertension | 0.903 (0.672-1.213) | 0.497 | 0.734 (0.386-1.398) | 0.347 | 0.975 (0.692-1.373) | 0.886 |
| Obstructive Sleep Apnea | 1.465 (0.590-3.644) | 0.411 | 0.647 (0.122-3.428) | 0.609 | 1.958 (0.654-5.859) | 0.23 |
| Obesity | 0.863 (0.525-1.419) | 0.563 | 2.411 (0.944-6.160) | 0.066 | 0.553 (0.294-1.039) | 0.066 |
| Diabetes | 1.043 (0.757-1.436) | 0.799 | 1.188 (0.602-2.344) | 0.619 | 0.949 (0.652-1.381) | 0.785 |
| Chronic Kidney Disease | 1.006 (0.665-1.522) | 0.978 | 0.975 (0.448-2.125) | 0.95 | 1.033 (0.623-1.713) | 0.9 |
| HIV | 2.460 (0.978-6.178) | 0.056 | 1.354 (0.141-12.975) | 0.793 | 2.729 (0.942-7.901) | 0.064 |
| Cancer | 1.335 (0.914-1.952) | 0.135 | 2.113 (0.962-4.637) | 0.063 | 1.068 (0.681-1.675) | 0.775 |
| Coronary Artery Disease | 1.323 (0.902-1.939) | 0.151 | 1.891 (0.774-4.618) | 0.162 | 1.203 (0.779-1.861) | 0.404 |
| Atrial Fibrillation | 0.874 (0.553-1.381) | 0.564 | 1.112 (0.369-3.353) | 0.851 | 0.860 (0.512-1.445) | 0.569 |
| Heart Failure | 0.892 (0.537-1.480) | 0.659 | 0.638 (0.219-1.857) | 0.409 | 0.928 (0.511-1.685) | 0.806 |
| Liver Disease | 1.275 (0.610-2.667) | 0.519 | 2.330 (0.406-13.370) | 0.343 | 1.307 (0.579-2.954) | 0.519 |
| Acute Kidney Injury | 1.067 (0.672-1.692) | 0.783 | 0.473 (0.194-1.153) | 0.099 | 1.619 (0.909-2.883) | 0.102 |
| **Labs** |  |  |  |  |  |  |
| WBC | 1.009 (0.985-1.033) | 0.468 | 0.992 (0.942-1.045) | 0.765 | 1.015 (0.988-1.044) | 0.279 |
| Creatinine > 1.2 | 1.659 (1.261-2.181) | <0.001 | 1.980 (1.099-3.564) | 0.023 | 1.584 (1.155-2.173) | 0.004 |
| Anion Gap | 1.087 (1.053-1.121) | <0.001 | 1.060 (1.003-1.121) | 0.04 | 1.102 (1.058-1.147) | <0.001 |
| Potassium | 1.177 (1.018-1.361) | 0.027 | 1.053 (0.813-1.363) | 0.695 | 1.236 (1.033-1.480) | 0.021 |
| ALT | 1.001 (0.999-1.002) | 0.358 | 1.001 (0.998-1.005) | 0.397 | 1.000 (0.999-1.002) | 0.585 |
| **Hospital Conditions** |  |  |  |  |  |  |
| ICU | 4.255 (3.149-5.749) | <0.001 | 9.393 (4.802-18.394) | <0.001 | 3.397 (2.404-4.807) | <0.001 |
| Duration (days) | 0.967 (0.950-0.984) | <0.001 | 0.972 (0.937-1.009) | 0.141 | 0.964 (0.945-0.983) | <0.001 |

Abbreviations: ACE-I, angiotensin-converting enzyme; ARBs, angiotensin II receptor blockers; BMI, body mass index; BP, blood pressure; SBP, systolic blood pressure; DBP, diastolic blood pressure; COPD, chronic obstructive pulmonary disease; HIV, human immunodeficient virus; WBC, white blood cell count; ALT, alanine aminotransferase

We additionally evaluated ACE-I use and ARB use separately and across different exposure periods (anytime use, in-hospital use and outpatient use). ARB in-hospital use was significantly associated with mortality in the entire population (OR, 0.560; 95% CI, 0.371-0.846; P=0.006), the AA population (OR, 0.196; 95% CI, 0.074-0.516; P=0.001), but not the non-AA population (OR, 0.687; 95% CI, 0.427-1.106; P=0.122) which might be due to limited statistical power (1-β= 0.55 in the Wald-based power analysis) (Figure 2); while ACE-I use was not significantly associated with mortality in any cohort nor any drug exposure period (Figure S1). Again, the interaction term of ARB use and AA population is not significant (OR, 0.488; 95%CI, 0/185-1.292); P=0.149), but the direction of the effect suggests AA population who received ARB might have smaller odds (OR, 0.421) of in-hospital death compared to non-AA population (Table S2). Outpatient use was not significantly associated with in-hospital mortality for either medication class. We confirmed all the results with inverse propensity-weighted logistic regression (Table S5).

**Figure 2.**
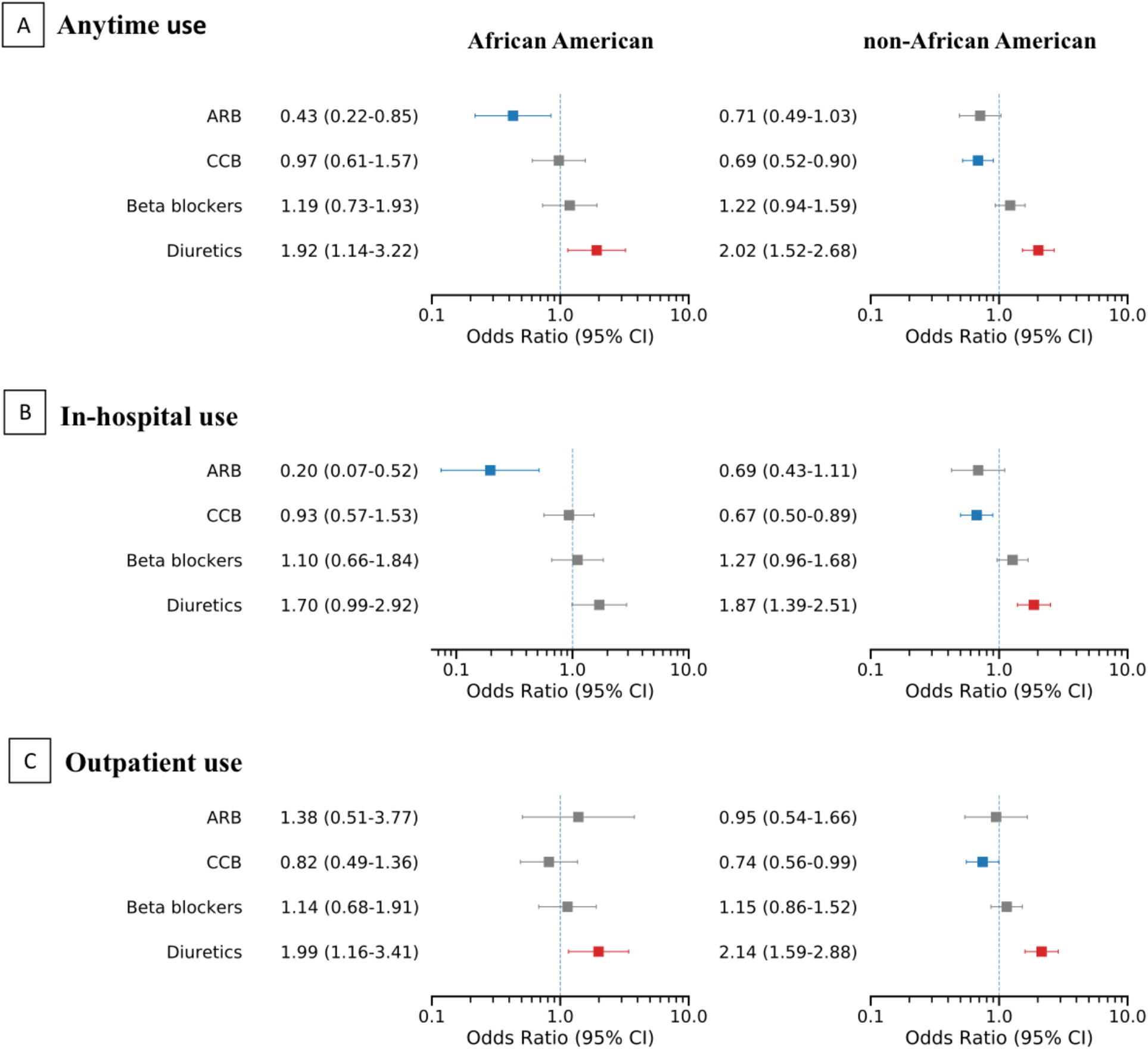
Different ARBs exposure times and other antihypertensive drugs, and their associations with COVID-19 in-hospital mortality. Abbreviations: ARB, angiotensin receptor blocker; CCB, calcium channel blocker

## Discussion

In this retrospective study of a racially and ethnically diverse cohort from New York City comprising 6,218 hospitalized COVID-19 patients seen between February 24 and March 31, 2020, we found that ACE-I/ARB use was significantly associated with reduced in-hospital mortality, particularly among African American (AA) patients. ACE-I/ARB users were older and had more comorbidities than non-users, but after controlling for these adverse risk factors, in-hospital ACE-I/ARB use (as opposed to outpatient-only use) was independently associated with improved outcomes. When analyzing ACE-I and ARB exposures separately, in-hospital ARB use was significantly associated with improved outcomes among AA patients but not non-AA patients. Conversely, ACE-I exposure was not independently associated with outcomes in any cohort.

Early in the COVID-19 pandemic, the finding that SARS-CoV-2 enters cells by binding to the *ACE2* protein led to concern that ACE-Is and ARBs might impact SARS-CoV-2 infectivity or virulence.^21^ Many clinical studies from the USA, Europe, and China were subsequently reported, examining the association between ACE-I/ARB use and COVID-19 outcomes. ^22–24^ These studies found that ACE-I/ARB usage was associated with neither harm nor benefit in the setting of COVID-19 in Caucasian population, however, one meta-analysis grouped studies by geography and found a lower mortality risk associated with ACE-I/ARB use among studies from China (OR = 0.65; 95% CI, 0.46−0.91; P = 0.013),^22^ which might be suggested by a higher ACE I/D allele frequency in Asian population.^13,25–27^ Whereas there was no significant association among studies from the US or Europe, suggesting the need to account for race and ethnicity. To our knowledge, this is the first study to report a potential benefit from the use of ACE-I/ARB in AA population hospitalized with COVID-19 using the one of the largest EMR systems in NYC.

Race and ethnicity may be relevant in this context as a proxy for genetic ancestry and the probability of carrying the *ACE* D-allele—the deletion allele of the *ACE* insertion/deletion polymorphism—which is associated with increased ACE1 activity and may contribute to ACE1/ACE2 imbalance in SARS-CoV-2 infection. Epidemiological studies in European, North African, and Asian settings have found significant positive correlations between D-allele prevalence and COVID-19 infection and mortality rates at the population level.^8–11,27^ While it is not practical at present to determine the *ACE* genotype of large cohorts of COVID-19 patients, the prevalence of the D-allele is known to vary geographically and is highest in Africa and the Middle East.^28^ Therefore, race may serve as a proxy for probability of D-allele carrier status. Indeed, we recently reported that African Americans have a higher prevalence of the D-allele than white Americans.^13^

With this premise in mind, we investigated the interaction between AA race and ACE-I/ARB use in relation to inpatient COVID-19 outcomes. Individuals carrying the D-allele would be expected to have excessive ACE1 and angiotensin-II in the context of COVID-19 infection and may thus derive benefit from either ACE1 inhibition by ACE-Is, or angiotensin-II blockade via ARB.^13^ Using a multi-ethnic cohort of 6,218 hospitalized COVID-19 patients, we found a significant association between ACE-I/ARB use and reduced in-hospital mortality overall; this association appeared to be driven by a strong protective association between ACE-I/ARB use and in-hospital mortality in the AA (but not the non-AA) population though the interaction between ACE-I/ARB use and race was not significant. While our ability to detect a significant interaction may have been limited by sample size, these findings are consistent with the hypothesis that RAS hyperactivity in an *ACE* D-allele-enriched (e.g., African American) COVID-19 positive cohort may benefit from ACE-I/ARB use.^13^

We further analyzed ACE-I or ARB use separately and according to either outpatient or in-hospital exposure. Neither in-hospital nor outpatient ACE-I use was associated with mortality. Conversely, in-hospital ARB use was significantly associated with reduced mortality among AAs but not non-AAs. This may suggest that ACE1 inhibition alone is inadequate in the context of ACE2 occupation/down-regulation by SARS-CoV-2 virions; excess angiotensin-II may still accumulate in the absence of sufficient ACE2 activity. Inhibiting downstream angiotensin-II type 1 receptors (AT1R) using ARB may therefore be more effective. Mechanistic and clinical studies to dissect the relationships between COVID-19, ACE1/ACE2 activity, angiotensin-II levels, and ARB are warranted. Attention should be paid to underlying *ACE* genotype in studies of RAS inhibitors in COVID-19 moving forward. The ongoing randomized clinical trial of ramipril against placebo to treat COVID-19 patients (NCT04366050) may provide insightful data on the therapeutic value of ACE inhibitors.^29^

Prior clinical studies in US population have examined associations between ACE-I/ARB use and COVID-19 outcomes without stratification by race.^16,30–32^ Most of these studies have found no association between ACE-I/ARB use and outcomes such as hospitalization, ICU admission, or in-hospital mortality. However, these cohorts had a substantially higher proportion of Caucasian (45-70%) patients than ours (23.3%). One exception was the study by Lam et al., which found lower rates of ICU admission and mortality among patients who continued rather than discontinued ACE-I/ARB treatment upon admission for COVID-19.^30^ The Lam et al. cohort was predominantly Caucasian (61.2%) but there was a large fraction of patients of “Other ethnicity” (26.2%), which could have influenced the overall D-allele prevalence in the cohort. Taken together, these results suggest that race and ethnicity—while imperfect as surrogates of D-allele prevalence—should be accounted for when assessing ACE-I/ARB associations with COVID-19.

### Limitations

Our study has several limitations. It was a retrospective study and cannot establish a causal relationship between ACE-I/ARB and COVID-19 outcomes via the *ACE* D-allele without individual-level germline genetic data and prospective treatment assignment. Secondly, our study is based on a single center experience and these analyses should be repeated in other cohorts. Finally, while outpatient medication adherence could not be verified, this EMR dataset draws on multiple data sources to bolster accuracy and comprehensiveness.

## Conclusions

In conclusion, our results agree with current recommendations to continue ACE-I/ARB in hospitalized COVID-19 patients provided there are no other contraindications. Our results also suggest a potential benefit for ACE-I/ARB among African American patients with COVID-19. We hypothesize that this is due to a higher *ACE* D-allele prevalence in the African American population. Further investigations in COVID-19 cohorts with linked clinical and genomic data are warranted.

## Data Availability

The Mount Sinai Health System database is not publicly available. We utilized Python and R, and their open-source libraries to conduct our analysis tailored to MSH data. The code is available upon request.

## Acknowledgements

The authors would like to acknowledge Sema4 IT for computational support hosted on AWS.

## Author contributions

Designed the study: LL and EES; Performed data acquisition and quality control: SL, ZW, ES; Performed statistical analysis: SL; Contributed clinical interpretation: TJ, RS, MAK, EG, NRN, RC, EES, LL; Drafted the paper: SL, RS, TJ, LL; Revised and approved the paper: MAK, EG, NRN, RC, EES.

## Supplementary Materials

**Figure S1.**
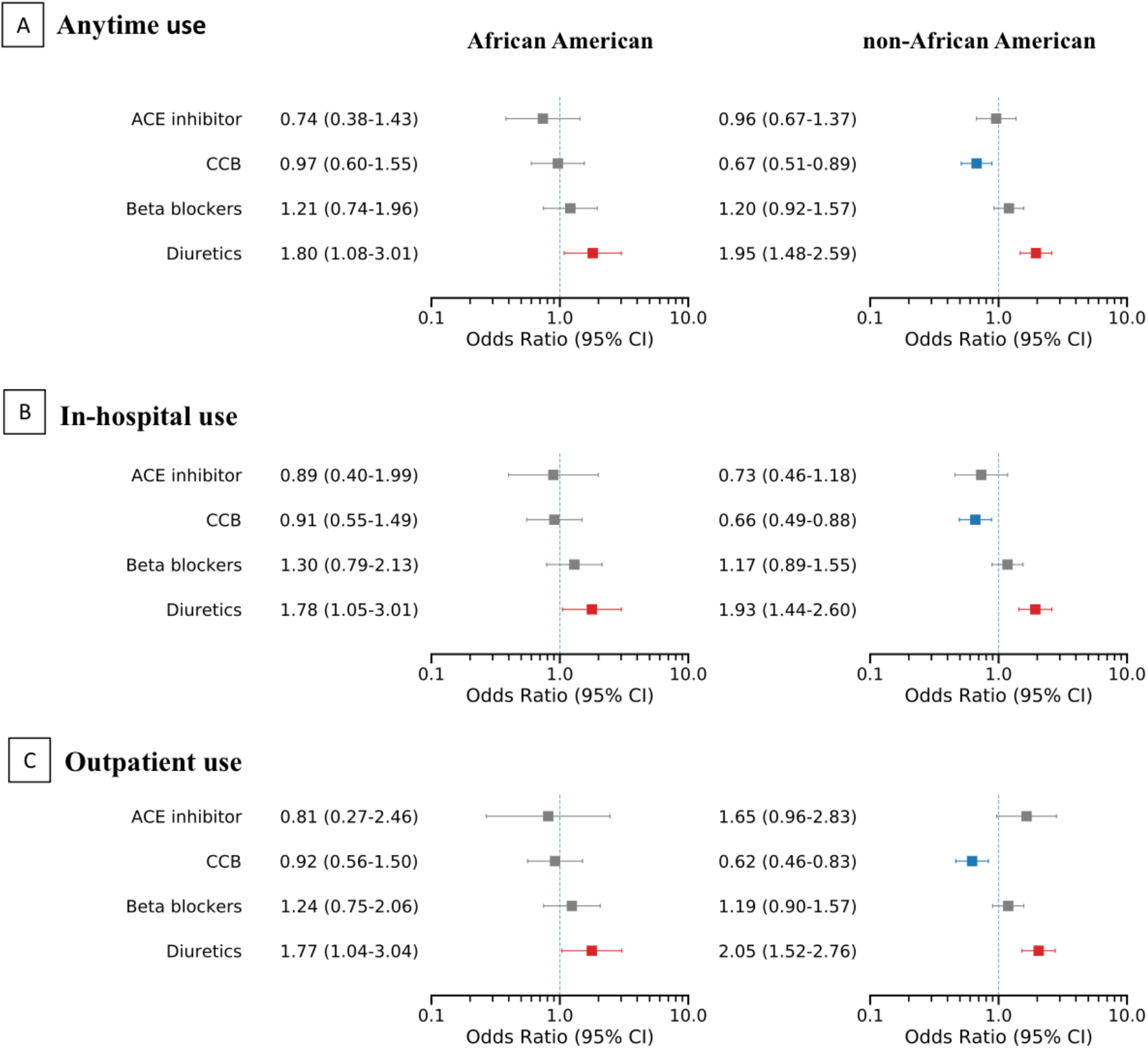
Different ACE-I exposure times and other antihypertensive drugs, and their associations with COVID-19 in-hospital mortality. Abbreviations: ACE, angiotensin-converting enzyme; CCB, calcium channel blocker

**Table S1.** ACE-I/ARB and potential confounding medication categories including detailed medication names.

| **Medication Category** | **Medication Names** |
| --- | --- |
| ACE inhibitors | Benazepril hydrochloride, Captopril, Enalapril maleate, Fosinopril sodium, Lisinopril, Moexipril, Perindopril, Quinapril hydrochloride, Ramipril, Trandolapril |
| ARBs | Azilsartan, Candesartan, Eprosartan mesylate, Irbesartan, losartan potassium, Olmesartan, Telmisartan, Valsartan |
| Diuretics | Chlorthalidone, Chlorothiazide, Hydrochlorothiazide, Indapamide, Metolazone, Amiloride hydrochloride, Spironolactone, Triamterene, Eplerenone, Furosemide, Bumetanide, Torsemide, Ethacrynic acid |
| Beta Blockers | Acebutolol, Atenolol, Betaxolol, Bisoprolol fumarate, Carteolol hydrochloride, Metoprolol, Nadolol, Nebivolol, Penbutolol sulfate, Pindolol, Propranolol hydrochloride, Solotol hydrochloride, Timolol maleate |
| Calcium-channel Blockers | Amlodipine, Bepridil, Diltiazem hydrochloride, Felodipine, Isradipine, Nicardipine, Nifedipine, Nisoldipine, Verapamil hydrochloride |

**Table S2.** Interaction between ACE-I/ARB and African American population from multivariable logistic regression also adjusted by other confounders.

| Interaction Terms | Anytime use |  | In-hospital use |  | Outpatient use |  |
| --- | --- | --- | --- | --- | --- | --- |
|  | Adjusted OR (95% CI) | p-value | Adjusted OR (95% CI) | p-value | Adjusted OR (95% CI) | p-value |
| ***ACE-I/ARB use*** |  |  |  |  |  |  |
| ACE-I/ARB | 0.708 (0.528-0.949) | 0.021 | 0.605 (0.420-0.871) | 0.007 | 1.122 (0.726-1.732) | 0.605 |
| ACE-I/ARB * AA | 0.736 (0.428-1.266) | 0.268 | 0.825 (0.413-1.648) | 0.585 | 0.695 (0.311-1.550) | 0.373 |
| AA | 0.896 (0.672-1.193) | 0.451 | 0.897 (0.673-1.198) | 0.463 | 0.879 (0.660-1.172) | 0.381 |
| ***ACE-I use*** |  |  |  |  |  |  |
| ACE-I | 0.908 (0.640-1.287) | 0.588 | 0.662 (0.414-1.058) | 0.085 | 1.661 (0.983-2.807) | 0.058 |
| ACE-I * AA | 0.817 (0.413-1.617) | 0.562 | 1.375 (0.572-3.303) | 0.476 | 0.490 (0.168-1.426) | 0.191 |
| AA | 0.844 (0.648-1.101) | 0.212 | 0.836 (0.640-1.092) | 0.188 | 0.832 (0.638-1.085) | 0.176 |
| ***ARB use*** |  |  |  |  |  |  |
| ARB | 0.708 (0.493-1.016) | 0.061 | 0.669 (0.419-1.068) | 0.092 | 0.998 (0.581-1.712) | 0.993 |
| ARB * AA | 0.797 (0.403-1.577) | 0.516 | 0.488 (0.185-1.292) | 0.149 | 1.299 (0.477-3.540) | 0.609 |
| AA | 0.854 (0.654-1.114) | 0.244 | 0.863 (0.661-1.126) | 0.278 | 0.842 (0.645-1.099) | 0.206 |

**Note:** among patients of anytime use, the odds ratio of in-hospital death in AA population who received ACE-I/ARB compared to non-AA population who also received ACE-I/ARB is calculated as $0.736 \times 0.896 = 0.659$ ; among patients of in-hospital use, the odds ratio of in hospital-death in AA population who received ARB compared to non-AA population who also received ARB is calculated as $0.488 \times 0.863 = 0.421$

**Table S3.** Characteristics of COVID-19 patients in African American and non-Africana American population.

|  | **African American** | **Non-African American** | **P-Value** |
| --- | --- | --- | --- |
| **n** | **1631** | **4584** |  |
| **Antihypertensive Medications** |  |  |  |
| ACE-I/ARB, n (%) | 314 (19.3) | 825 (18.0) | 0.277 |
| ACE inhibitor, n (%) | 161 (9.9) | 444 (9.7) | 0.866 |
| ARB, n (%) | 171 (10.5) | 426 (9.3) | 0.176 |
| Calcium-channel blockers, n (%) | 464 (28.4) | 1085 (23.7) | <0.001 |
| Beta blockers, n (%) | 403 (24.7) | 1066 (23.3) | 0.249 |
| Diuretics, n (%) | 422 (25.9) | 1179 (25.7) | 0.929 |
| **Patient Characteristics** |  |  |  |
| Age, median [Q1,Q3] | 61 [47,72] | 62 [46,75] | 0.012 |
| Sex, n (%) |  |  | <0.001 |
| Female | 854 (52.4) | 2015 (44.0) |  |
| Male | 777 (47.6) | 2569 (56.0) |  |
| BMI (kg/m<sup>2</sup>), mean (SD) | 30.2 (8.7) | 28.3 (7.1) | <0.001 |
| Smoking status, n (%) |  |  | <0.001 |
| Never | 859 (52.7) | 2514 (54.8) |  |
| Not Asked | 330 (20.2) | 1063 (23.2) |  |
| Quit | 339 (20.8) | 844 (18.4) |  |
| Yes | 103 (6.3) | 163 (3.6) |  |
| Temperature (F), mean (SD) | 98.9 (1.6) | 99.0 (1.8) | 0.02 |
| Max. temperature (F), mean (SD) | 100.2 (2.0) | 100.5 (2.0) | <0.001 |
| O2 saturation (%), median [Q1,Q3] | 97.0 [95.0,99.0] | 96.0 [93.0,98.0] | <0.001 |
| Min. O2 saturation (%), median [Q1,Q3] | 92.0 [87.0,97.0] | 91.0 [84.0,96.0] | <0.001 |
| Heart rate (BPM), mean (SD) | 95.9 (18.4) | 95.0 (20.2) | 0.116 |
| Respiratory rate >25, n (%) | 142 (8.7) | 596 (13.0) | <0.001 |
| High BP (SBP>140 or DBP>90), n (%) | 633 (38.8) | 1446 (31.5) | <0.001 |
| **Comorbidities** |  |  |  |
| Asthma, n (%) | 90 (5.5) | 194 (4.2) | 0.039 |
| COPD, n (%) | 65 (4.0) | 121 (2.6) | 0.008 |
| Hypertension, n (%) | 466 (28.6) | 1152 (25.1) | 0.007 |
| Obstructive Sleep Apnea, n (%) | 37 (2.3) | 69 (1.5) | 0.053 |
| Obesity, n (%) | 145 (8.9) | 291 (6.3) | 0.001 |
| Diabetes, n (%) | 297 (18.2) | 735 (16.0) | 0.047 |
| Chronic Kidney Disease, n (%) | 188 (11.5) | 351 (7.7) | <0.001 |
| HIV, n (%) | 35 (2.1) | 49 (1.1) | 0.002 |
| Cancer, n (%) | 115 (7.1) | 319 (7.0) | 0.945 |
| Coronary Artery Disease, n (%) | 117 (7.2) | 447 (9.8) | 0.002 |
| Atrial Fibrillation, n (%) | 65 (4.0) | 237 (5.2) | 0.065 |
| Heart Failure, n (%) | 101 (6.2) | 238 (5.2) | 0.143 |
| Chronic Viral Hepatitis, n (%) | 24 (1.5) | 30 (0.7) | 0.004 |
| Liver Disease, n (%) | 23 (1.4) | 93 (2.0) | 0.139 |
| Acute Kidney Injury, n (%) | 108 (6.6) | 232 (5.1) | 0.021 |
| **Labs** |  |  |  |
| WBC (K/uL), median [Q1,Q3] | 7.3 [5.3,10.0] | 7.7 [5.6,10.8] | 0.001 |
| Creatinine > 1.2, n (%) | 603 (37.0) | 1258 (27.4) | <0.001 |
| Anion Gap (mEq/L), mean (SD) | 13.4 (4.9) | 12.8 (4.0) | <0.001 |
| Potassium (mmol/L), mean (SD) | 4.4 (0.9) | 4.4 (0.8) | 0.023 |
| ALT (U/L), median [Q1,Q3] | 25.0 [17.0,45.0] | 30.0 [19.0,53.0] | <0.001 |
| **Hospital Conditions** |  |  |  |
| ICU, n (%) | 222 (13.6) | 763 (16.6) | 0.004 |
| Days after infection, median [Q1,Q3] | 3.8 [0.6,8.5] | 3.8 [0.7,8.6] | 0.407 |
| Hospital status, n (%) |  |  | 0.003 |
| Censored | 144 (8.8) | 364 (7.9) |  |
| Deceased | 300 (18.4) | 1026 (22.4) |  |
| Discharged | 1187 (72.8) | 3194 (69.7) |  |

**Table S4.** Adjusted odds ratios of secondary outcomes for African American COVID-19 patients associated with ACE-I/ARB and other covariates.

|  | ICU (N=1,487) |  | AKI (N=1,487) |  |
| --- | --- | --- | --- | --- |
|  | Adjusted OR (95% CI) | p-value | Adjusted OR (95% CI) | p-value |
| **Antihypertensive Medications** |  |  |  |  |
| ACE-I/ARB | 0.709 (0.419-1.198) | 0.198 | 1.306 (0.721-2.364) | 0.379 |
| Calcium-channel blockers | 0.953 (0.599-1.514) | 0.838 | 1.043 (0.617-1.762) | 0.876 |
| Beta blockers | 1.986 (1.253-3.146) | 0.003 | 1.671 (0.989-2.824) | 0.055 |
| Diuretics | 4.728 (2.918-7.660) | <0.001 | 0.590 (0.325-1.071) | 0.083 |
| **Patient Characteristics** |  |  |  |  |
| Age | 0.985 (0.969-1.002) | 0.082 | 0.994 (0.974-1.015) | 0.563 |
| Male | 1.102 (0.692-1.755) | 0.682 | 0.954 (0.557-1.631) | 0.862 |
| Smoking Status: Not Asked | 2.437 (1.362-4.360) | 0.003 | 1.173 (0.597-2.307) | 0.644 |
| Smoking Status: Quit | 1.020 (0.582-1.787) | 0.946 | 0.763 (0.402-1.448) | 0.407 |
| Smoking Status: Yes | 2.595 (1.127-5.976) | 0.025 | 0.776 (0.261-2.313) | 0.649 |
| BMI | 1.034 (1.008-1.060) | 0.011 | 0.988 (0.956-1.020) | 0.455 |
| Temperature | 0.755 (0.655-0.872) | <0.001 | 0.910 (0.759-1.090) | 0.307 |
| Max. Temperature | 1.510 (1.310-1.740) | <0.001 | 0.857 (0.716-1.026) | 0.093 |
| O2 Saturation | 0.996 (0.963-1.029) | 0.793 | 0.976 (0.941-1.013) | 0.206 |
| Min. O2 Saturation | 0.970 (0.960-0.981) | <0.001 | 0.999 (0.984-1.014) | 0.877 |
| Heart Rate | 1.009 (0.997-1.021) | 0.163 | 1.001 (0.988-1.014) | 0.939 |
| Respiratory Rate > 25 | 2.138 (1.156-3.954) | 0.015 | 1.070 (0.518-2.211) | 0.855 |
| High BP (SBP>140 or DBP>90) | 0.891 (0.567-1.399) | 0.615 | 0.735 (0.431-1.253) | 0.257 |
| **Comorbidities** |  |  |  |  |
| Asthma | 0.891 (0.352-2.257) | 0.808 | 3.210 (1.212-8.499) | 0.019 |
| COPD | 0.557 (0.201-1.544) | 0.261 | 1.517 (0.400-5.742) | 0.540 |
| Hypertension | 0.873 (0.488-1.563) | 0.649 | 1.675 (0.867-3.237) | 0.125 |
| Obstructive Sleep Apnea | 2.457 (0.848-7.121) | 0.098 | 0.211 (0.022-1.986) | 0.174 |
| Obesity | 0.468 (0.214-1.025) | 0.058 | 1.491 (0.611-3.641) | 0.380 |
| Diabetes | 1.513 (0.873-2.622) | 0.140 | 0.644 (0.345-1.200) | 0.166 |
| Chronic Kidney Disease | 0.371 (0.179-0.770) | 0.008 | 0.858 (0.443-1.663) | 0.650 |
| HIV | 1.963 (0.537-7.173) | 0.308 | 0.918 (0.217-3.879) | 0.907 |
| Cancer | 1.054 (0.494-2.249) | 0.891 | 0.801 (0.307-2.089) | 0.650 |
| Coronary Artery Disease | 1.733 (0.808-3.713) | 0.158 | 0.511 (0.199-1.315) | 0.164 |
| Atrial Fibrillation | 0.628 (0.229-1.723) | 0.367 | 1.558 (0.580-4.183) | 0.379 |
| Heart Failure | 1.486 (0.650-3.399) | 0.348 | 0.449 (0.150-1.346) | 0.153 |
| Liver Disease | 0.291 (0.043-1.983) | 0.207 | 1.907 (0.352-10.332) | 0.454 |
| **Labs** |  |  |  |  |
| WBC | 1.034 (0.991-1.078) | 0.119 | 1.034 (0.983-1.088) | 0.192 |
| Creatinine > 1.2 | 0.685 (0.414-1.133) | 0.141 | 38.538 (11.297-131.468) | <0.001 |
| Anion Gap | 1.044 (0.999-1.091) | 0.053 | 1.045 (1.001-1.091) | 0.044 |
| Potassium | 1.132 (0.881-1.455) | 0.334 | 1.059 (0.831-1.351) | 0.642 |
| ALT | 0.999 (0.996-1.002) | 0.573 | 1.004 (1.000-1.007) | 0.044 |
| **Hospital Conditions** |  |  |  |  |
| Duration (days) | 1.053 (1.019-1.087) | 0.002 | 1.007 (0.967-1.049) | 0.723 |

**Table S5.** Inverse propensity-weighted logistic regression results for COVID-19 in-hospital mortality in different race groups and ACE-I/ARB exposure time.

| **Drug (Year 2019-2020)** | **All Races** |  | **African American** |  | **Non-African American** |  |
| --- | --- | --- | --- | --- | --- | --- |
|  | **Adjusted OR (95% CI)** | **p-value** | **Adjusted OR (95% CI)** | **p-value** | **Adjusted OR (95% CI)** | **p-value** |
| **ACE-I/ARB use** |  |  |  |  |  |  |
| Anytime use | 0.673 (0.511-0.886) | 0.005 | 0.381 (0.204-0.712) | 0.003 | 0.765 (0.556-1.052) | 0.100 |
| In-hospital use | 0.577 (0.418-0.795) | < 0.001 | 0.307 (0.140-0.672) | 0.003 | 0.653 (0.450-0.948) | 0.025 |
| Outpatient use | 0.941 (0.626-1.415) | 0.770 | 0.534 (0.224-1.273) | 0.157 | 0.927 (0.567-1.518) | 0.764 |
| **ACE-I use** |  |  |  |  |  |  |
| Anytime use | 0.951 (0.689-1.313) | 0.761 | 0.684 (0.321-1.458) | 0.326 | 1.016 (0.708-1.456) | 0.933 |
| In-hospital use | 0.724 (0.489-1.071) | 0.106 | 4.113 (0.409-41.37) | 0.230 | 0.787 (0.496-1.250) | 0.311 |
| Outpatient use | 1.347 (0.846-2.144) | 0.209 | 0.824 (0.192-3.526) | 0.794 | 1.730 (0.970-3.086) | 0.063 |
| **ARB use** |  |  |  |  |  |  |
| Anytime use | 0.636 (0.464-0.870) | 0.005 | 0.428 (0.190-0.966) | 0.041 | 0.655 (0.454-0.946) | 0.024 |
| In-hospital use | 0.514 (0.340-0.775) | 0.002 | 0.144 (0.049-0.428) | < 0.001 | 0.609 (0.384-0.966) | 0.035 |
| Outpatient use | 0.761 (0.480-1.209) | 0.248 | 0.637 (0.187-2.166) | 0.470 | 0.594 (0.351-1.005) | 0.053 |

